# Transcranial photobiomodulation influences BOLD responses during finger sequence execution: An fMRI Study in young and older adults

**DOI:** 10.64898/2026.07.06.26357423

**Authors:** Marjorie Dole, Vincent Auboiroux, Daniel Anglade, Emilie Cousin, Monica Baciu, Caroline Sandre-Ballester, Sandrine Rebecchi, Tigrane Cantat-Moltrecht, John Mitrofanis

## Abstract

Transcranial photobiomodulation (PBM) is an emerging non-invasive brain stimulation method that is thought to increase neural metabolism by stimulating ATP production by the mitochondria. However, the mechanisms of action and the effects on the human brain are still unclear. In the present study, we investigated the potential of this method to enhance Blood Oxygen Level Dependent (BOLD) responses during the execution of a motor task in young and aged participants. Sixty young and aged participants were included in this single-blinded, sham-controlled, randomised, crossover study. They underwent an fMRI recording before and after 24-min stimulation with a 80-LEDs helmet emitting transcranially red and near infrared light. Post *vs* Pre BOLD signal was compared between PBM and SHAM, in each group. At baseline, aged participants showed reduced BOLD signal compared to young ones, in key regions of the sensorimotor processing, principally the left primary motor cortex and striatum. Transcranial PBM did not have a real impact in the young group. In aged participants, analysis performed on the whole brain did not reveal widespread changes, but rather local increases in BOLD responses in the right ventral premotor cortex and insula. More specifically, a regional analysis further showed increased BOLD responses in the left primary motor cortex, and right dorsal and ventral premotor regions and striatum. Some of these regions were under-activated at baseline in aged participants relatively to the young ones. These results suggest that transcranial PBM can increase fMRI BOLD responses locally in some task-related regions, particularly in aged subjects. Further research are needed to distinguish neural from vascular effects in transcranial PBM.

## 1. INTRODUCTION

In our ever-increasing aging world, finding a way to offset or even slow the deleterious effects of age on cognitive function, together with helping the brain to improve resilience mechanisms, is a major challenge. In this context, non-invasive neurostimulation techniques such as transmagnetic or transcranial electric stimulation have become attractive options for both clinicians and scientists to stimulate brain regions and improve brain function, including promoting brain plasticity, increasing functional communication between brain regions and potentially improving cognitive performances in several pathological conditions (Koch et al. 2024). These techniques have proven effective in acting on neural function and present minimal risks comparatively to invasive techniques. However, they are limited in several ways, for example, difficulties in focusing the stimulation for transcranial electric stimulation, or difficulties to reach deep regions of the brain. Furthermore, most studies to date focus on single target regions, despite the fact that the functions being targeted typically rely on brain activity distributed across networks rather than a unique region (Toth et al. 2024).

Recently, another method, involving transcranial application of red and near infrared light, has been shown to stimulate neural metabolism and promote better neural functioning. This method, known as transcranial photobiomodulation (PBM), acts mainly at the level of the mitochondria, improving energy production in the cells through increased adenosine triphosphate (ATP) production. This feature is of particular interest to the process of aging, because mitochondrial dysfunction is central in the generation of oxidative stress and the cellular abnormalities observed associated with aging (Dhote et al. 2021). At the cellular level, PBM is thought to act through a number of photoacceptors, the best known being cytochrome c oxidase, located in the chain IV of the mitochondria. The net result is an increased cytochrome c oxidase activity, mitochondrial membrane potential and ATP production in both cell cultures (Hoh Kam and Mitrofanis 2023; Wong-Riley et al. 2005) and animal models (Salehpour et al. 2017; Cardoso et al. 2022a). In humans, a recent pilot study using ^31^P-MR Spectroscopy has reported an improved rate of ATP synthase flux after one-week stimulation in seven older subjects (Fear et al. 2023) suggesting an enhanced cell metabolism. Increased cytochrome c oxidase concentration and increased oxyhaemoglobin/deoxyhaemoglobin ratio have also been reported consistently using near infrared spectroscopy techniques, for both young (Wang et al. 2016; Wang et al. 2017) and older participants; for the latter, the effect has appeared even more marked (Saucedo et al. 2021). In fact, it is generally accepted that PBM has a greater impact on systems (cell cultures and animal models) suffering from pathology and/or oxidative stress than on those that are healthy and functioning normally (Banqueri et al. 2019; Cardoso et al. 2022b). For these key reasons, PBM is of particular interest for therapeutic and preventive purposes in ageing and disease (Dole and Mitrofanis, 2025). In addition, it has been proposed that the interaction between photons and cytochrome c oxidase would result in a nitric oxide dissociation from the binuclear centre of the cytochrome c oxidase, resulting in increased levels of circulating nitric oxide. As a well-known vasodilator, nitric oxide promotes increased cerebral blood flow near the illuminated region (Uozomi et al. 2010; Salgado et al. 2015), potentially enhancing oxygen supply to neurons. Quite remarkably, these positive outcomes have been reported, not only within the illuminated region, but also across other brain regions, presumably through the activation of functionally connected networks or systemic effects (Dmochowski et al. 2020; Yang et al. 2025; Johnstone et al. 2014).

A key issue that has limited the widespread use and acceptance of transcranial PBM as a therapeutic tool has been the scarcity of large-scale, placebo controlled clinical studies (Dole et al. 2023; Salehpour et al. 2019). Recent efforts have been made toward this purpose, principally in healthy volunteers, to clarify the mechanisms of action and characterise the effects of transcranial PBM on the brain. For example, a fMRI study from Dmochowski et al. (2020) found that after a one-session PBM of the left dorsolateral prefrontal cortex in twenty young heathy participants, there was an increased functional connectivity between this region and the rest of the brain. More recently, a randomised study in 55 aged participants has reported increased functional connectivity in the frontoparietal network after a 12-min stimulation of the left dorsolateral prefrontal cortex; this was associated with an increase in the performances in a working memory test (Yang et al. 2025). There are also reports of a change in cortical oscillations at rest, particularly in the alpha and beta frequency ranges (Wang et al. 2021; Pruitt et al. 2024). However, the precise impact of transcranial PBM on brain activity during the performance of a task and how this is modulated by age is still unclear. A decrease in activation has been found in parietal and frontal areas during a working memory task, after seven daily sessions of PBM; however, the study in question did not have a control group and therefore, practice effects cannot be ruled out (Hu et al. 2023). fMRI studies using finger tapping tasks have also suggested modifications of the BOLD (blood oxygen level dependent) signal in task-related brain regions, the primary motor cortex and putamen, following transcranial PBM (Bibb et al. 2025; El Khoury et al. 2019). However, the pattern of change differed across studies; for example, one study reported a decrease BOLD response in the left primary motor cortex and putamen (El Khoury et al. 2019) whereas another showed an increase in the same regions (Bibb et al. 2025). One possible reason for these discrepancies could be in the devices and parameters used (stimulation localisation, wavelength, energy applied, stimulation time, pulsing, or pulse frequency), aspects that have much confounded the PBM field over the years (Salehpour et al. 2018; Fernandes et al. 2024).

In the present study, we aimed to investigate the effect of transcranial PBM in both young and aged subjects during the performance of a motor sequences task. We used a sequential finger mobilisation paradigm, where subjects had to execute finger sequences in response to visual stimuli. This task is known to engage a left-lateralised network comprising the primary motor, premotor and supplementary motor areas, dorsolateral prefrontal cortex, superior/inferior parietal cortex, striatum and cerebellum (Witt et al. 2008; Daselaar et al. 2003). Based on evidence of age-related alterations in cortico-striatal circuits, we hypothesised baseline differences in task-related BOLD responses between young and older subjects. Specifically, we expected reduced engagement of the striatum in older participants (Seidler et al. 2010; Fitzroy et al. 2020), while no strong directional prediction was made for primary sensorimotor regions given the dependence of the age-related effects on task complexity (Van Ruitenbeek et al. 2023). Aged participants were also expected to engage a more extended network with greater involvement of homologous ipsilateral regions, frontal and parieto-occipital areas (Seidler et al. 2010; Turesky et al. 2016). We further hypothesise that transcranial PBM will change task-related BOLD responses in sensorimotor regions, (Bibb et al. 2025), particularly in those regions showing age-related differences.

## 2. MATERIAL AND METHODS

This study (NCT05845216) was conducted in the dedicated research laboratory (CEA-Leti Clinatec) at Grenoble University Hospital (CHU Grenoble Alpes) between September 2023 and December 2024. It was approved by the Clinical Committee Review Board (CPP Est-II, approval number ID-RCB: 2023-A00507-38). Informed consent was obtained from all participants prior to enrolment, and the study was conducted in accordance with the Declaration of Helsinki, Good Clinical Practice guidelines, and applicable national laws and regulations.

### 2.1. Participants

Sixty right-handed subjects were enrolled: 30 in the young group and 30 in the aged group. Participants were recruited via local advertisements. They were first pre-screened by phone call to confirm handedness and the absence of contra-indication to the MRI exam. During the inclusion visit, they were interviewed regarding their medical history and current treatments and eligibility criteria were verified. If eligible, they signed an informed consent form and a neuropsychological assessment was done (Montreal Cognitive Assessment, MoCA; Nasreddine et al. 2005) to exclude cognitive deficits.

The inclusion criteria were as following: 1) 18-40 year olds (young group) and 60-85 year olds (aged group); 2) right handedness (Edinburgh Handedness Inventory score >70%; Oldfield, 1971); 3) MoCA score >26; 4) normal or corrected-to-normal vision. Exclusion criteria were: 1) contra-indication to MRI; 2) contra-indication to PBM; 3) neurological or psychiatric disorder; 4) current use of antidepressant or antipsychotic medication; 5) known motor impairment.

One participant of the young group was excluded due to an experimental error, two participants of the aged group were excluded because of the discovery of cerebral anomalies observed on the T1-weighted MRI, and one additional participant in the aged group was excluded as a behavioural outlier (>3 SD from the group mean). Analyses were therefore conducted on 29 participants in the young group (mean age: 29.55; s.d.: 7.06) and 27 participants in the aged group (mean age 66.15 year old; s.d. 4.63). The demographic information collected from both groups is shown on Table 1.

**Table 1.** Demographic characteristics of participants. ***: statistical difference on a student t-test p<0.001.

|  | **young (n=29)** | **aged (n=27)** | **p-value** |
| --- | --- | --- | --- |
|  | *Mean (S.D.)* | *Mean (S.D.)* |  |
| **Age** | 29.55 (7.06) | 66.15 (4.63) | <.0001*** |
| **Handedness** | 94.62 (8.88) | 96.68 (8.23) | 0.37 |
| **Gender** | 14 F<br>15 H | 14 F<br>13 H | NA |
| **Phototype** | 2.97 (0.86) | 2.96 (0.85) | 0.99 |
| **MoCA score** | 28.79 (1.11) | 28.22 (1.22) | 0.073 |
| **Randomisation** |  |  |  |
| *PBM / Sham* | 15 | 14 |  |
| *Sham / PBM* | 14 | 13 |  |

The participants differed only in their age group, but not on their handedness, phototype or MoCA score (Table 1). The randomisation was balanced, with half participants in the PBM/SHAM order and half participants in the SHAM/PBM order in both groups.

### 2.2. Experimental Protocol

#### 2.2.1. Study design

This study was a prospective, single-blinded, sham-controlled, crossover trial involving two groups of participants, an ‘aged’ group and a ‘young’ group. Participants completed two visits: one with an ‘Active’ stimulation (PBM), and one with a ‘Sham’ stimulation (SHAM). The order of the stimulation was randomised between participants (PBM/SHAM or SHAM/PBM), and the randomisation was made through an electronic case report form (e-CRF). Randomisation was stratified by gender, ensuring an equal number of male and female participants in both groups. To account for the unknown duration of subsequent after-effects of the PBM treatment, a wash-out period of 4 weeks (+/- 2 weeks) was implemented between visits.

To ensure blinding, the PBM and SHAM stimulation devices were identical in appearance; the SHAM was, in effect, the same device, but light emission was blocked with black adhesive tape. In addition, participants wore an eye mask over their eyes for the entire stimulation session. Spectrophotometer measurements were performed prior to the study to confirm that the light output from the SHAM device was negligible.

The study followed a Pre-Post design: at each visit, participants underwent fMRI scanning before (‘Pre’ session) and after (‘Post’ session) the 24-min treatment (PBM or SHAM). As the PBM device was not MRI compatible, the treatment was administered outside the scanner.

#### 2.2.2. Task-fMRI

During each of the four MRI sessions (prePBM, postPBM, preSHAM, postSHAM), participants laid supine in the scanner for approximately 40 minutes per session. Visual instructions were displayed on a screen located behind the scanner, and viewed by participants via a mirror mounted in front of their eyes. The motor task was executed using a 5-button response box placed over the right hand.

The task involved executing finger-movement sequences during sixteen 20-s blocks. Instructions about the sequence to execute were presented visually as 5-digit sequences, with each digit corresponding to a specific finger. Participants were instructed to repeat the sequence continuously and as quickly as possible, without pausing during the entire block. Motor block were interleaved with 20-s rest blocks during which participants fixated sequences of five “#” symbols and remained still. Individual button presses were recorded. Prior to scanning, participants completed a 10-min training session to familiarise themselves with the procedure, followed by a brief in-scanner training consisting in two sequences immediately before the task.

MRI images were acquired on a 1.5T Siemens Espree scanner with a 12-channels head coil. The scanning sequence order was as following: motor task-fMRI, two resting-states runs, T1-weighted anatomical MRI, B0 field map. For the task-related fMRI, a T2* weighted echo planar imaging (EPI) sequence sensitive to blood oxygen level dependent (BOLD) contrast was used, provided by the Centre for Magnetic Resonance Research (CMRR) (Moeller et al. 2010, Feinberg et al. 2010, Xu et al. 2013), with a multiband factor of 2. Acquisition parameters were as following: repetition time (TR) 1.67s, echo time (TE) 50ms, flip angle 90°, field of view (FOV) 224mm. Thirty-four axial slices covering the entire cerebrum and parallel to the anterior-posterior commissure plane were acquired in an interleaved order. The reconstructed voxel size was 3 x 3 x 3.5 mm. A total of 480 volumes were acquired, corresponding to a 13-min acquisition duration. One T1-weighted anatomical MRI was also acquired for each session (MPRAGE sequence, 4 per participants), which were merged during the preprocessings. The parameters for this acquisition were: sagittal acquisition, TR = 3000ms, TE = 2.65ms, flip angle = 8°, FOV = 256mm, isotropic 1mm^3^ voxel size.

While this study was part of a larger project on the effect of PBM on brain activity in young and aged adults, only task-fMRI results are reported here.

#### 2.2.3. Photobiomodulation procedure

After each ‘Pre’ fMRI session, participants were comfortably seated in a quiet room to receive their assigned treatment (PBM or SHAM). To maintain blinding, participants wore a black eye-mask and were instructed to remain still and relaxed throughout the 24-min session.

The PBM device used in this study was the commercially available WellRed Coronet Duo ® (https://wellred.com.au/). The device was a helmet comprising 40 LEDs at 670nm and 40 LEDs at 810nm designed to illuminate the entire head and delivered pulsed light with a pulse frequency 40Hz and a duty cycle respectively at 60% and 50%. Each wavelength was administered for 12 minutes, resulting in a total stimulation time of 24 minutes. The optical power output of each LED at a 100% duty cycle was measured at 345mW for the 670-nm LEDs and 267mW for 810-nm LEDs. Consequently, the total energy applied after the 12 minutes session was thus 207J per 670-nm LED and 96J for the 810nm LEDs.

At the end of the PBM session, participants returned to the MRI scanner where they underwent a second session, identical to the first one.

#### 2.2.4. Data analysis

##### 2.2.4.1. Behavioural analyses

R studio (https://www.R-project.org) was used for the analyses of the behavioural data. Individual button presses recorded in the scanner were used to compute the mean number of taps per block (Nb_taps). For each subject and treatment, the Post-Pre difference was calculated. Normality of data distribution was assessed using a Shapiro-Wilk test. As the data were not distributed normally, the effect of the treatment on the Post-Pre difference was evaluated in each group using a signed-rank Wilcoxon test.

##### 2.2.4.2. fMRI data analysis

The first three volumes of each scanning session, during which the fMRI signal reaches a steady-state, were discarded from the analyses. Images were first arranged to follow the BIDS (Brain Image Data Structure) format (Gorgolewski et al. 2016, a, b). Preprocessing was performed using *fMRIPrep* 23.2.0 (Esteban et al. 2019; Esteban et al. 2018). Briefly, structural preprocessing implied averaging T1w images across sessions, bias field correction, brain tissue segmentation, and spatial normalisation to the MNI152 standard space. Functional preprocessings included susceptibility distortion correction, slice time correction, correction for head motions and registration to the anatomical volume. Finally, realigned and normalised functional images were smoothed with an 8-mm FWHM kernel. More detailed preprocessing procedures can be find in *fmriprep* citation boilerplate in supplementary materials S1.

Statistical analyses were performed using SPM12 (Statistical Parametric Mapping; Wellcome Department of Cognitive Neurology, UK, https://www.fil.ion.ucl.ac.uk/spm/software/spm12) running on Matlab R2023b (The Mathworks Inc., Natick, USA). The AAL3 toolbox (Automatized Anatomical Labelling; Rolls et al. 2020), and, when necessary, the Yale BioImage Suite (mni2tal mapping from Lacadie et al., 2008, http://www.bioimagesuite.org) were used for effect localisation.

Statistical analysis was performed using the General Linear Model (GLM). At the first level, BOLD responses for the ‘Task’ blocks were modelled using a boxcar function (20-s duration) convolved with the canonical haemodynamic response function (HRF). Six motion parameters, together with global signal, CSF and white matter signals were added in the model as nuisance regressors. Number of taps was added as a covariate. One contrast image representing task (vs. implicit baseline) was generated per session (prePBM, postPBM, preSHAM, postSHAM) for each participant and subsequently entered in the second-level analysis.

At the second level, a random-effect analysis (RFX) was performed to investigate the effects of Timing (Pre, Post) and Treatment (PBM, SHAM) on task-related brain responses in each group. A full factorial model was implemented, with factors including Age (young, aged), Timing (Pre, Post) and Treatment (PBM, SHAM). Unless otherwise stated, the statistical threshold for parametric maps was set at p<0.05, family-wise error (FWE) corrected at the peak-level. For exploratory purposes, results at p<0.001 uncorrected (peak-level) with a cluster extent k≥30 voxels are also reported.

Analyses were performed in three steps: (1) examination of brain activations elicited by task performance in each group; (2) comparison between groups at baseline, that is, difference between aged and young in the mean of both ‘Pre’ sessions; (3) investigation of the effect of the treatment in each group, that is, the difference (Post-Pre)_PBM_-(Post-Pre)_SHAM_.

Given our interest in sensorimotor regions, we performed both a whole-brain analysis and a region of interest (ROIs) analysis restricted to nine regions known to be involved in finger movement tasks, namely left and right primary motor cortex, left and right dorsal and ventral premotor cortices, left and right putamen and supplementary motor area (Witt et al. 2008). These ROIs were obtained from an activation map derived from a Neurosynth meta-analysis of 83 studies on finger movements (https://neurosynth.org; keyword: “finger movements”; date: 26/8/2025). The generated activation map was first thresholded using a false discovery rate (FDR) approach at p<0.01 (height threshold z=4). To separate these clusters into independent regions, we combined these clusters with the Human Motor Area Template (HMAT; Mayka et al. 2006) using Marsbar (Brett et al. 2002), retaining only activated voxels overlapping with HMAT regions. The resulting nine ROIs are shown on Figure 5A. In these regions, the Marsbar toolbox was used to extract mean contrast estimates and the contrast (Post-Pre)_PBM_-(Post-Pre)_SHAM_ in each age group was tested. Results were considered as statistically significant for a p<0.05, FDR-corrected for multiple comparisons.

#### 2.2.5. Assessment of the blinding

At the end of the study, in order to check the effectiveness of the blinding process, we asked the participants if they could guess the order in which they received the two treatments. On the 56 participants assessed, 29 guessed the correct order, and 27 seven did not. A binomial test confirmed that this pattern (51.78%) did not significantly differ from chance level (50%, p=0.89, 95% CI = [38%-65%]).

#### 2.2.6. Estimating the illuminated regions: Monte Carlo simulations

The quantification of light energy reaching the cortex is a complex question in the photobiomodulation field as light propagation is complicated to measure in vivo in the human brain. In order to quantify the amount of energy reaching our regions of interest, we used Monte Carlo simulation using an open-source MMC simulator (Fang and Yan, 2019). Detailed procedure for these stimulations and a map representing the estimated light diffusion with our device can be found in Supplementary Material S2.

## 3. RESULTS

### 3.1. Behavioural results

The effect of treatment (PBM, SHAM) on the Post-Pre difference in the mean number of taps per block was assessed in each group. Figure 1 shows that there was an increase in the number of taps for both treatments in Post vs Pre sessions; this difference tended to be larger for PBM than for SHAM in the Young group even if this did not reach statistical significance (Md PBM = 3.43, Md SHAM = 2.69, W=296, p=0.092, effect size r=0.25, small). In the Aged group, no difference was observed between PBM and SHAM treatments (Md PBM: 1.22, Md Sham = 1.61, W= 145, p=0.296).

**Figure 1.**
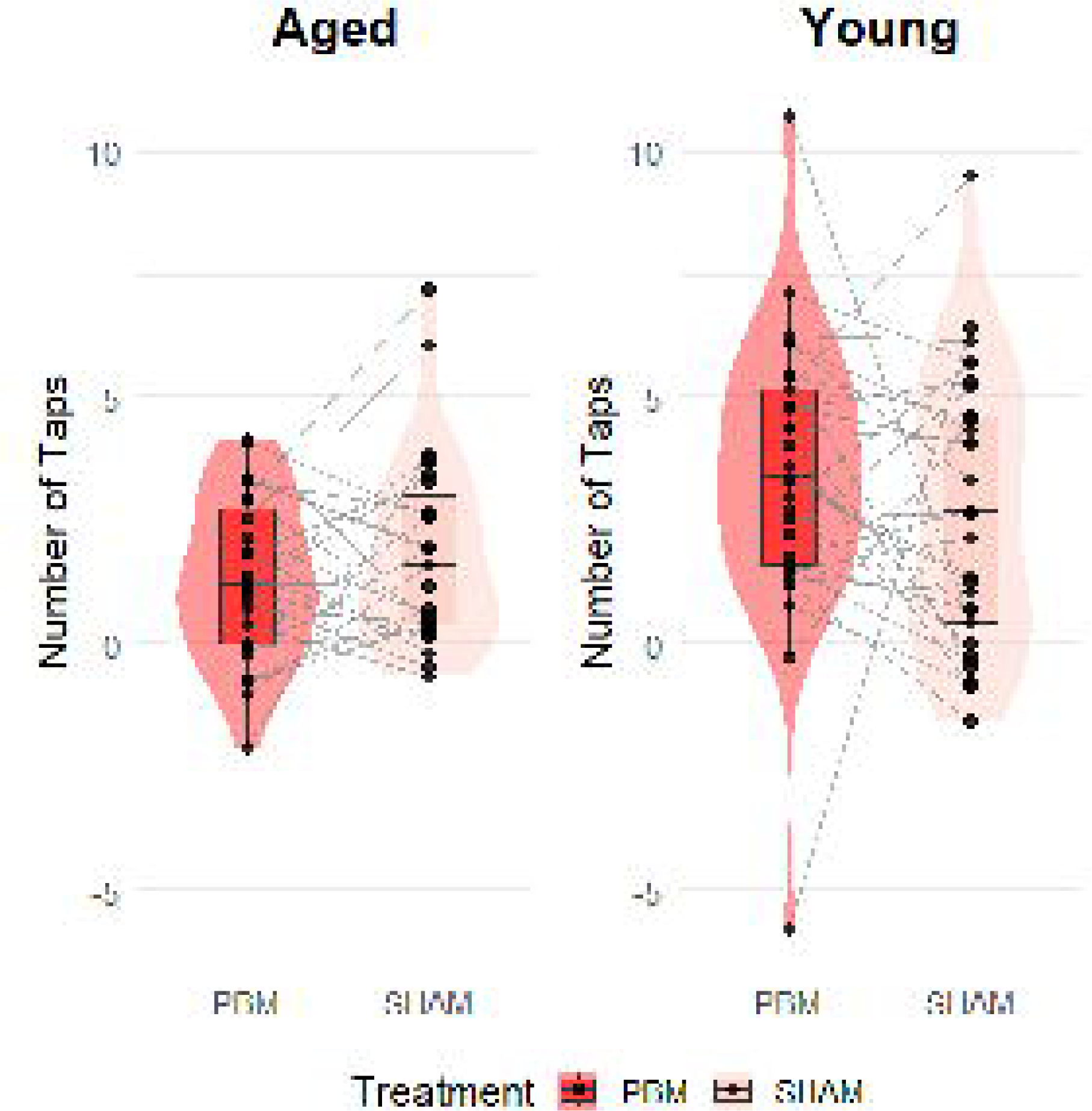
Post-Pre differences in number of taps per block as function of treatment (PBM, SHAM) and age group (Young, Aged)

### 3.2. fMRI Results

#### 3.2.1. Main effect of Task

In a first analysis, we explored the brain regions activated by the task in both young and aged subjects; the main effect of ‘Task’ was examined for each group separately. We found that, for both groups, the task execution involved a widespread network of regions located including primary sensori-motor regions (left M1/S1), the supplementary motor area (SMA), dorsal and ventral premotor areas (PMd/PMv), superior parietal lobule, the striatum, anterior insula and the right cerebellum (Figure 2).

**Figure 2.**
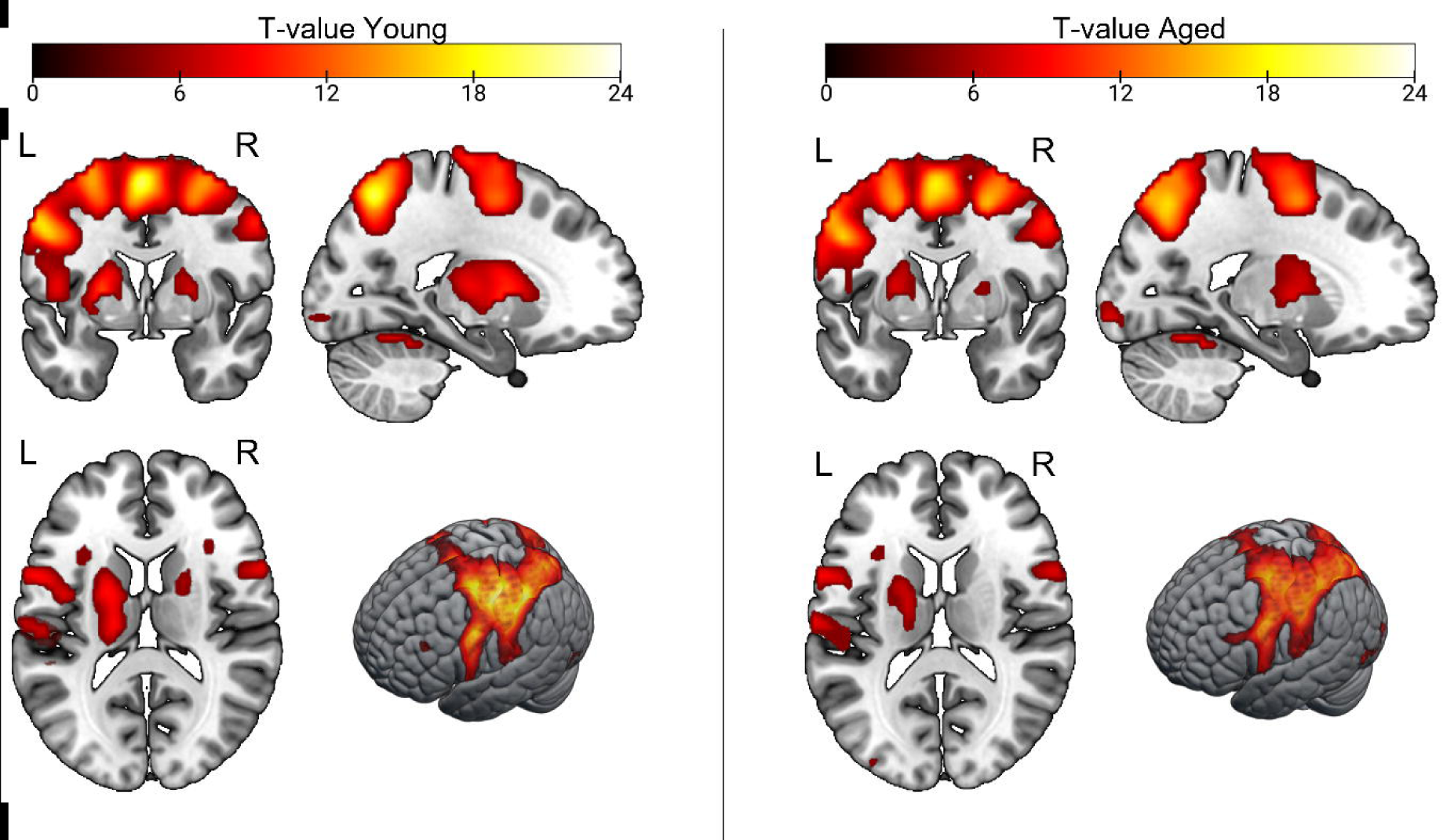
Brain activations evoked by the main ‘Task’ contrast, in young (left) and aged (right) subjects. Activations are overlayed on the MNI 152 Template. L=Left. R=Right. Maps are thresholded at a p<0.05 FWE-corrected at the peak-level.

#### 3.2.2. Group differences on brain activations before treatment

Next, we investigated whether the execution of the task elicited age-related differences in the patterns of activation. For this, we compared BOLD responses during the task between the two groups before treatment (young>aged and aged>young contrasts in both ‘Pre’ sessions). Figure 3 indicates that when contrasting young>aged, BOLD responses were reduced in aged subjects relative to the young ones in the left primary sensorimotor cortex (M1/S1) and the striatum, mainly in the left side, and in the right cerebellum. At a lower statistical threshold, we found additional clusters in the right striatum and insula. For the reverse contrast (aged>young), aged subjects showed an increased BOLD signal relative to the young ones in the superior parietal lobule/precuneus (left BA7), SMA/paracentral lobule and right primary motor cortex. At an exploratory statistical threshold (p<0.001 uncorrected, k≥30), additional clusters were detected in the left ventral premotor cortex and ventral parts of the primary motor cortex (Figure 3; Table 1).

**Figure 3.**
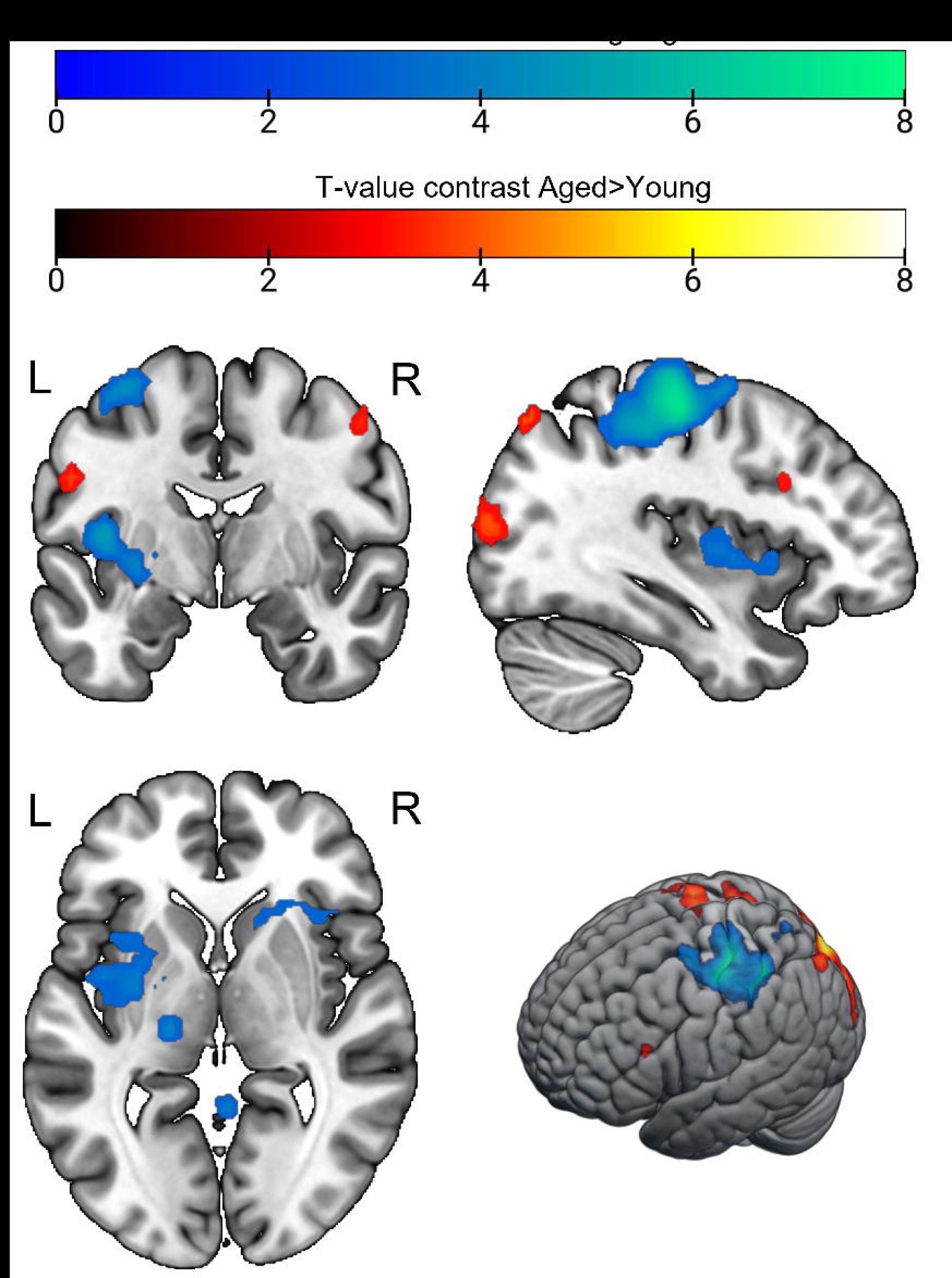
BOLD response maps for the contrast aged>young and young>aged in the ‘Pre’ sessions, overlayed on the MNI 152 template. For representation purpose, maps are thresholded at p<0.001 uncorrected at the voxel-level, with an extent threshold k=30 voxels. Corresponding FWE-corrected results can be found in Table 2. Cold colours represent the contrast young>aged, warm colours the contrast aged>young. L= Left. R=Right.

**Table 2.**
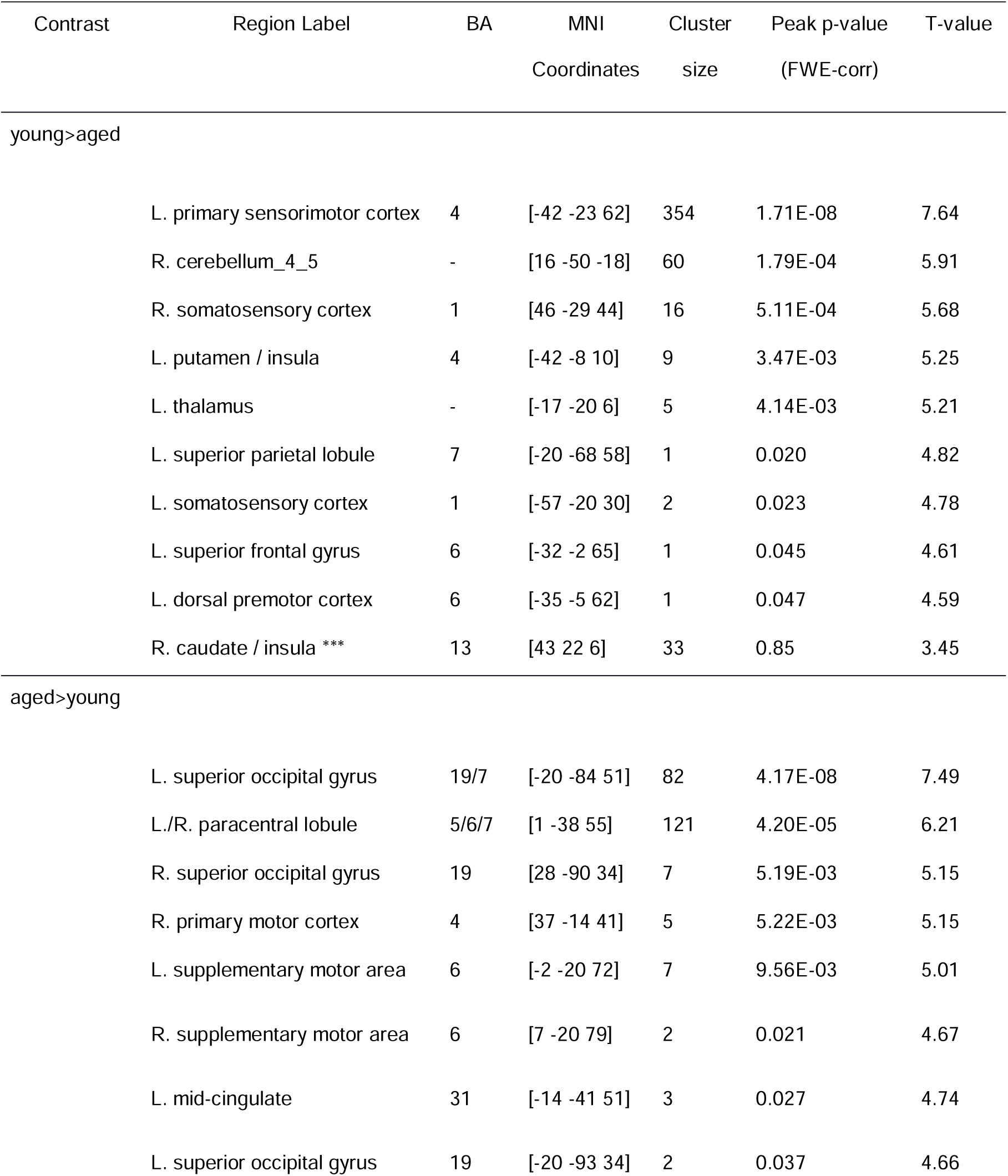
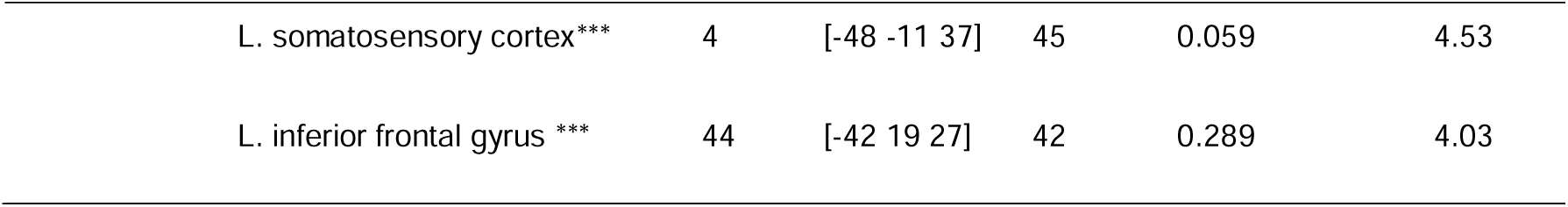
Peak localisation and statistics for young>aged and aged>young contrasts, thresholded at p<0.05 FWE-corrected at the peak level. ***: indicates significant peaks at a lower threshold, p<0.001 uncorrected (peak-level) with an extend threshold k≥30 voxels. L. = left, R. = right. BA: Broadman’s area.

In summary there were clear differences in the patterns of responses when undertaking the task in the aged group compared to the young group. For some regions, namely the left primary sensorimotor regions, mainly contralateral to the movement, the bilateral striatum and the right cerebellum, the aged group showed reduced responses relatively to the young group. Conversely, for other regions, the aged group showed increased BOLD responses in the right primary motor cortex, bilateral occipital gyrus and paracentral lobules extending into posterior supplementary motor area, relative to the young one.

#### 3.2.3. Effect of treatment. Post>Pre activations for PBM and SHAM: whole brain analysis

To investigate the effect of treatment on these patterns of responses, we first ran a whole-brain analysis of the Timing*Treatment interaction, that is, the contrast (Post-Pre)_PBM_-(Post-Pre)_SHAM_, in both groups.

In the aged group, there was a significant difference between PBM and SHAM in a region located over the right inferior frontal gyrus and insula (MNI coordinates [49 7 2]; p_FWE_ = 0.042, T-value=4.62, cluster size 40 voxels) when considering the (Post-Pre)_PBM_-(Post-Pre)_SHAM_ contrast. Further inspection revealed that BOLD signal increased in (Post-Pre)_PBM_ in the bilateral striatum and right ventral premotor cortex, whereas no such increase was observed for the SHAM treatment even at lower thresholds (Table 3). In the young group, direct testing of the interaction ((Post-Pre)_PBM_-(Post-Pre)_SHAM_ contrast) yielded no significant differences, even at a lower threshold. However examining the main contrasts separately showed that the (Post-Pre)_PBM_ contrast elicited increased BOLD response in the right inferior frontal gyrus / ventral premotor cortex and insula, whereas the (Post-Pre)_SHAM_ contrast revealed only marginal increase in the supramarginal gyrus and medial prefrontal cortex at the uncorrected threshold (Table 3 and Figure 4A).

**Figure 4.**
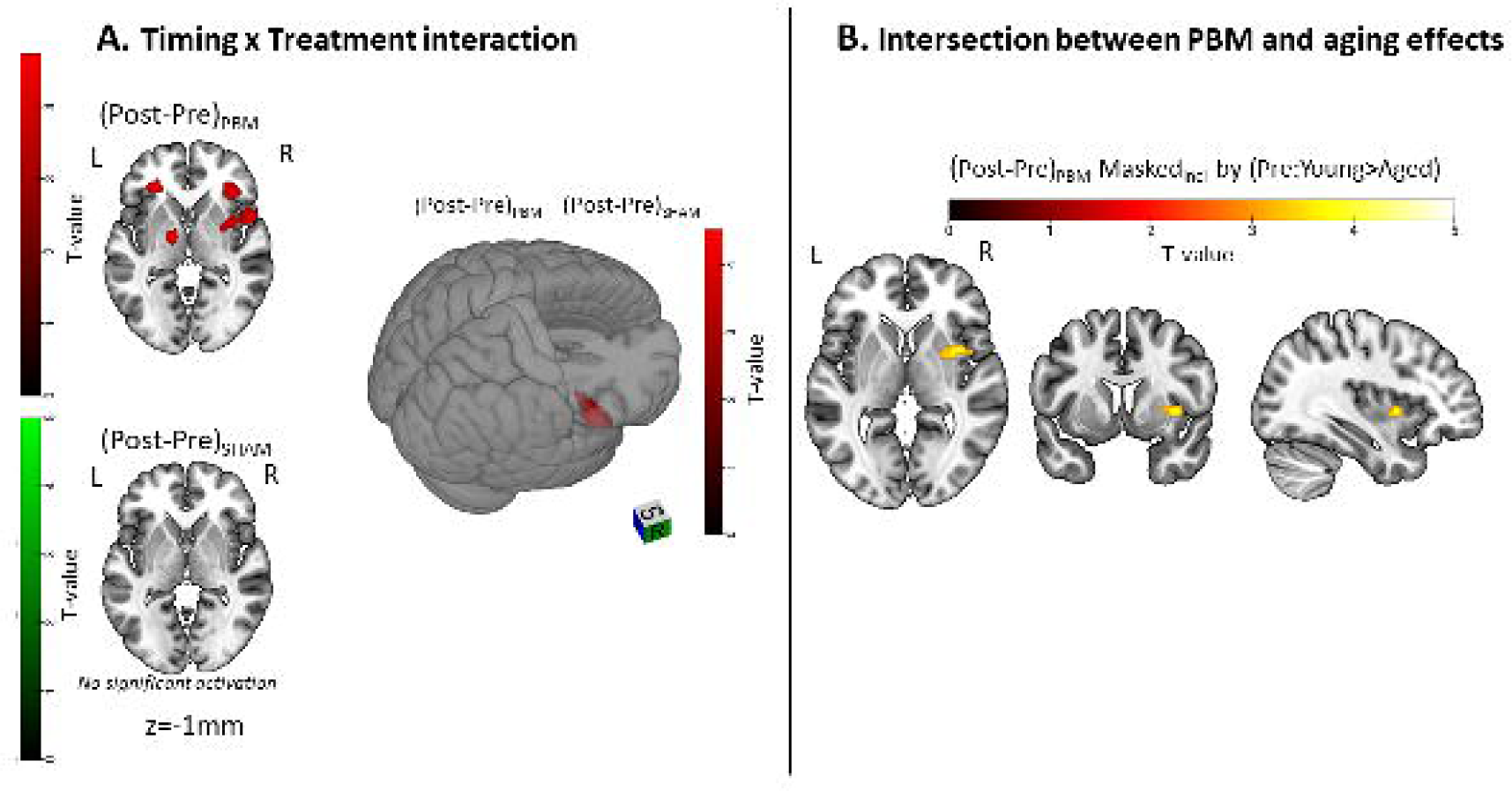
**A. Timing*Treatment interaction in the aged group**. BOLD responses resulting from the [(Post-Pre)_PBM_-(Post-Pre)_SHAM_] contrast are displayed on a 3D rendering of the MNI152 template. For representation purpose, maps were thresholded at an uncorrected threshold p<0.001 (peak level), k≥30 voxels. Corresponding FWE-corrected results can be found in Table 3. **B. Intersection between PBM effects and aging effects**. BOLD responses obtained for the (Post-Pre)_PBM_ contrast were masked inclusively (p_mask_<0.01) with the Pre:young>aged contrast. Activations are overlaid on the MNI152 template and thresholded with a p<0.001 uncorrected (peak-level), k≥30 voxels

**Table 3.** Localisation and statistics of increased responses in Post vs Pre sessions, for both treatments and in both groups. *** indicates significant peaks at a lower threshold, p<0.001 uncorrected (peak-level) with an extend threshold k=30 voxels.

| Contrast | Region Label | BA | MNI<br>Coordinates | Cluster<br>size | Peak p-value<br>(FWE-corr) | T-value |
| --- | --- | --- | --- | --- | --- | --- |
| aged: Post-Pre PBM |  |  |  |  |  |  |
|  | L. pallidum | - | [-17 -2 2] | 2 | 0.015 | 4.90 |
|  | L. thalamus | - | [-14 -8 -1] | 1 | 0.020 | 4.82 |
|  | R. putamen, inferior frontal gyrus | - /44 | [49 7 2] | 1 | 0.023 | 4.78 |
|  | L. inferior frontal gyrus | 47 | [-29 34 -1] | 2 | 0.024 | 4.77 |
|  | R. inferior frontal gyrus, insula *** | 45/47 | [34 31 -4] | 43 | 0.28 | 4.04 |
| aged: Post-Pre Sham |  |  |  |  |  |  |
|  | - | - | - | - | - | - |
| young: Post-Pre PBM |  |  |  |  |  |  |
|  | R. inferior frontal gyrus / insula | 44/13 | [49 16 -4] | 11 | 0.005 | 5.16 |
| young: Post-Pre Sham |  |  |  |  |  |  |
|  | L. supramarginal gyrus *** | 39/40 | [-69 -41 30] | 34 | 0.072 | 4.48 |
|  | L/R. Medial prefrontal cortex *** | 9 | [1 50 37] | 109 | 0.137 | 4.29 |

The reverse contrast, (Pre-Post)_PBM_ - (Pre-Post)_SHAM_ revealed no difference between PBM and SHAM, in neither group.

Then, we examined whether regions showing increased BOLD signal after PBM overlapped spatially with regions exhibiting reduced activity in the aged group compared with the young group at baseline. To this end, the contrast (Post>Pre)_PBM_ was tested in aged participants within an inclusive mask (p_mask_ < 0.01) defined by the contrast young>aged at baseline. This exploratory analysis, thresholded at p-<0.001 uncorrected (k≥30) indicated that the PBM-related increase in the right striatum/inferior frontal regions was located within areas showing reduced baseline activity in aged participants relative to young ones (T_peak_=4.01; coordinates [34 4 -1]; cluster extent 35 voxels; see Figure 4B). This exploratory result may be interpreted as a spatial convergence between PBM-related and age-related effects rather than confirmatory evidence of a mechanistic interaction.

#### 3.2.4. Effect of treatment. Post>Pre activations for PBM and SHAM: regional analysis

A main focus of this study was the effect of PBM in sensorimotor regions during the performance of the task. To do this, we also performed a regional analysis in those regions, by extracting mean contrast estimates in nine ROIs (Figure 5A), located in left and right primary motor cortex, dorsal and ventral premotor regions, putamen and supplementary motor areas. To examine treatment effects, we extracted mean contrast estimates for the [(Post-Pre)_PBM_-(Post-Pre)_SHAM_] contrast in each region and age group. To correct for multiple comparisons, an FDR-correction was applied, with differences considered as statistically significant p_FDR_<0.05. In aged participants, a significant Timing*Treatment interaction was observed, with larger (Post-Pre) increase for the PBM treatment in comparison with the SHAM, in the left primary motor cortex (t(26)=2.50, p_FDR_<0.05), right dorsal (t(26)=2.50, p_FDR_<0.05) and ventral (t(26)=2.70, p_FDR_<0.05) premotor cortices, right putamen (t(26)=2.28, p_FDR_<0.05) and supplementary motor area (t(26)=3.19, p_FDR_<0.05) indicating that Post-Pre difference was higher for PBM than for SHAM (see Figure 5B). The interaction failed to reach significance in the right primary motor cortex, left ventral premotor cortex, left putamen (all p_FDR_>0.05) and was only marginal in the left dorsal premotor cortex (t(26)=2.06, p_FDR_=0.06). Moreover, one-sample t-tests confirmed that the Post-Pre increase was significant for PBM in the left primary motor, left and right dorsal premotor cortex, left and right putamen and supplementary motor area (all p_FDR_<0.05), but not in the other regions. For SHAM, no significant increase occurred between Pre and Post sessions (all p_FDR_>0.05). In the young group, there was no effect of treatment, *ie*, no interaction Timing*Treatment observed in any of the ROIs.

**Figure 5.**
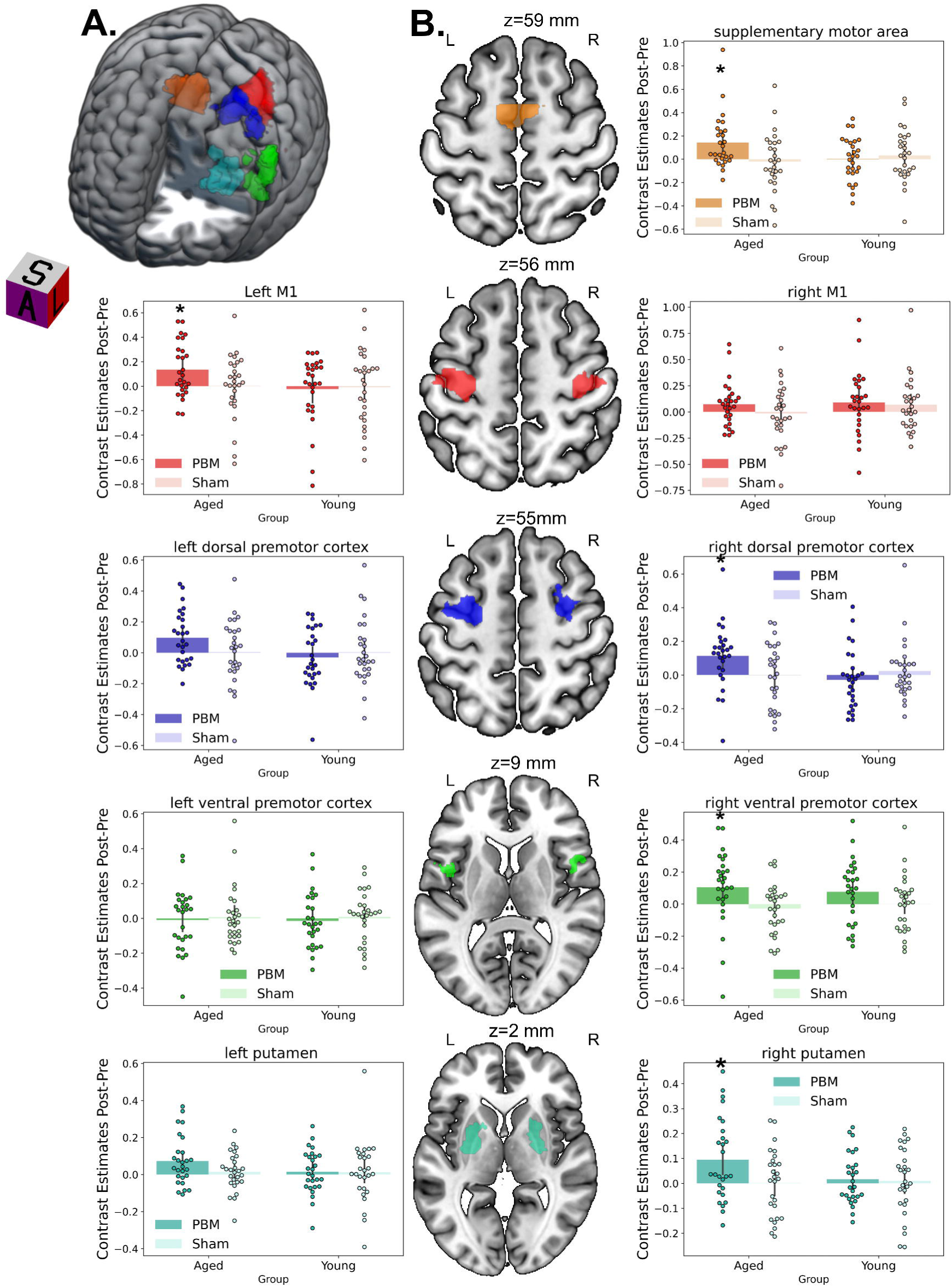
**A**. Example of the left-side regions of interest displayed on the MNI-152 template of the brain. Orange: supplementary motor area. Red: left primary motor cortex. Blue: left dorsal premotor cortex. Green: left ventral premotor cortex. Cyan: left striatum. **B.** (Post-Pre) contrast estimates extracted from the nine regions of interest in young and aged subjects for PBM and SHAM treatments. * indicates a significant (Post-Pre) difference (p_FDR_<0.05).

#### 3.2.5. Correlation with execution speed

Next, we investigated the correlation between (Post-Pre) change in BOLD responses and the (Post-Pre) change in movement execution speed (number of taps per block), in both groups. We also examined whether this correlation differed between treatments. As the motor execution speed was expected to correlate mainly with primary motor regions (Jäncke et al. 1998), analysis was restricted to the left and right primary motor cortex ROIs. In the young group, this analysis revealed a positive correlation between the (Post-Pre) signal change and the change in mean number of taps in the left primary motor cortex. This suggests that subjects who showed higher increase in the left primary motor cortex activity also showed higher increase in movement execution speed (Pearson correlation r=0.518, p<0.001). This relationship, however, did not appear to change between PBM and SHAM (Figure 6). In contrast, no significant correlation occurred in the aged group (Pearson correlation coefficient r=0.049, p>0.05). In the right primary motor cortex, no significant correlation was found between change in cerebral activity and change in execution speed in either group.

**Figure 6.**
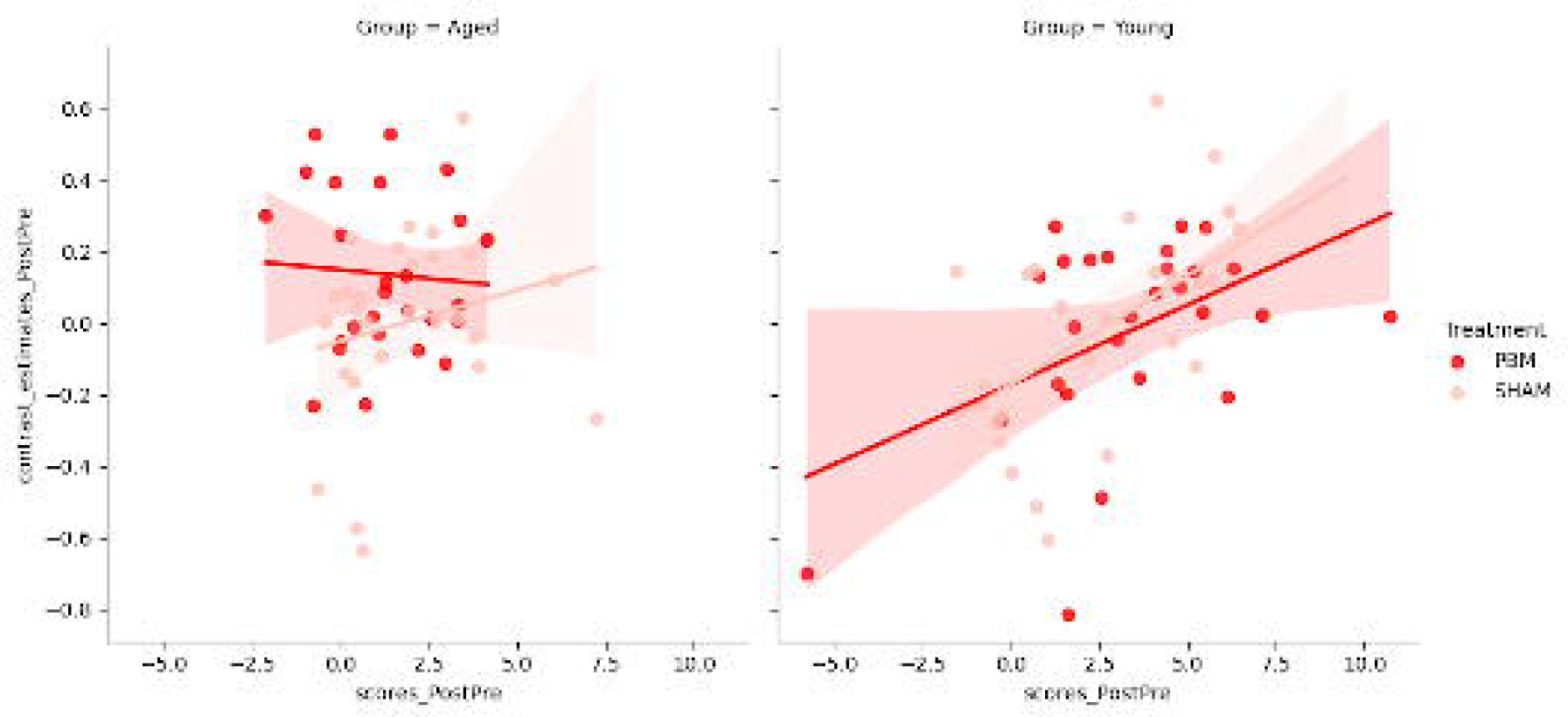
Correlation between Post-Pre change in BOLD in left primary motor cortex and change in execution speed, in the aged (left) and young (right) group, as function of the treatment.

#### 3.2.6. Evaluation of the persistence of the effect

Finally, in order to evaluate the persistence of tPBM effects throughout the session in aged subjects, each session (prePBM, postPBM, preSHAM, postSHAM) was divided in two equal halves. Analyses were then conducted to determine whether the tPBM-induced changes differed between early blocks (first half of the session) and late blocks (second half of the session). The results of this analysis are presented in supplementary material S3. Briefly, no substantial differences were observed between early and late blocks, suggesting that the tPBM-induced effects were maintained throughout the duration of the session.

#### 3.2.7. Assessment of potential motion-related confounds

In order to examine group differences in head motion and evaluate potential motion-related confounding effects, the mean framewise displacement (FD) was extracted for each subject and each session from the *fmriprep* outputs. The results of this analysis are presented in supplementary material S4.

Briefly, a significant effect of Age was observed, whereas no interaction with Timing and Treatment were detected. To determine whether the observed age-related differences could be explained by motion, baseline comparison between young and aged subjects were repeated with mean FD added as a covariable in the statistical model. As shown on supplementary material S4, the results remained largely unchanged, indicating that the reported effects were not substantially driven by differences in head motion.

## 4. DISCUSSION

The aim of the present study was to evaluate the ability of transcranial PBM to influence BOLD responses recorded by fMRI during the execution of sequences of finger movements. To investigate this, we conducted a randomised, sham-controlled, fMRI study with two groups of participants, an aged group and a young group. BOLD signal was recorded while participants performed the movements, both before and after a 24-min PBM or SHAM stimulation and Post-Pre changes were compared between PBM and SHAM conditions. Our main findings include that: (1) a single PBM session did not lead to behavioural changes (2) PBM induced distinct patterns of effects in young and aged brains and (3) exploratory analyses suggested a spatial overlap between regions showing PBM-related changes and those exhibiting reduced responses at baseline in aged subjects. These findings will be the main focus of discussion below. We will consider the transcranial penetration of light to the brain, and whether the mechanism of action of PBM reflects an underlying neural or vascular change.

### Transcranial penetration of light to the brain

One key question in PBM research is how much light actually reaches the cortex to induce a biological effect. As illustrated by supplementary figure S2 and Table S2, a small amount of applied energy did reach our functional regions of interest, principally the primary motor cortex, dorsal and ventral premotor areas and supplementary motor area. This suggests that even a small fraction of the initial light energy can trigger a biological response. It should be noted that we also obtained an increased BOLD signal in much deeper regions of the brain, namely the right striatum, where almost no light reached (∼10^-5^ Joules for the entire session). The basal ganglia are functionally strongly connected with motor regions and it has been shown that stimulating the left primary motor cortex using an electric or magnetic stimulation can also change activity in other related regions, such as the dorso-medial prefrontal cortex (Moisa et al. 2016), supplementary motor area, ventral premotor cortex and putamen (Wang et al. 2020). Other studies have also demonstrated that stimulating the left primary motor cortex changed its connectivity with the left and right putamen, or even between other regions linked to the sensorimotor network, for example between the thalamus and the right dorsal premotor cortex (Chen et al. 2022). There is thus a possibility that light-induced effects in superficial regions of the motor network propagate to other, often deeper, regions of the network even if they receive no light, as it has been already suggested for other neurostimulation methods (Beynel et al. 2020).

### Mechanism of action of PBM: neural or vascular?

An important consideration arising from these results relates to whether the PBM-induced increase in the BOLD signal reflects enhanced neural activity or primarily vascular effects. Although BOLD signal is tightly associated with neural activity, it is also the result of a complex mix of vascular and neural responses (Drew, 2019). PBM has been proposed to act both on regional cerebral blood flow through a NO-mediated vasodilation (Uozomi et al. 2010) and on neural cell function through a boost of ATP production and neural metabolism (Yan et al. 2025) and a change in neural excitability (Konstantinovic et al. 2013). The relative contribution of these components in the alteration of neurovascular coupling is still to be determined. Recently, some authors have suggested an effect driven primarily by endothelial cells and smooth muscle of the vessels (Truong et al. 2022); however, how this relates precisely with neural excitability and EEG findings having showed increased power in the alpha, beta and gamma frequency ranges is still to elucidate (Shadahdian et al. 2024). More research should be dedicated to this topic, for example with simultaneous recordings of fMRI/EEG or fMRI/ASL measurements. Moreover, PBM could also have an impact on astrocytes function, another component of the neurovascular coupling (El Massri et al. 2016). Our present results do not allow us to dissociate neural, vascular, and glial contributions and cannot entirely be solely attributed to neural mechanisms. However, the persistence of the effect throughout the scanning session (supplementary material S3), beginning several minutes after the end of stimulation and ending after ∼20 minutes, argues against a purely transient vascular effect and is more consistent with sustained neurometabolic or neurophysiological changes induced by PBM, even if vascular contributions cannot be fully excluded.

Concerning the haemodynamic consequences of tPBM, another additional comment can be done, regarding the duration of the effects. It is still unclear, how long a single session of PBM does affect the haemodynamic parameters. This could have impacted our results, as our PBM device was not MRI compatible and there was a small delay (approximately 5 to 8 min) between the end of the PBM session and the beginning of the subsequent MRI session. Near infrared spectroscopy studies, having investigated haemodynamic parameters during and immediately following a 8-min stimulation have showed a persistence of effects until 5 to 6 min after the stimulation (Wang et al. 2017; Saucedo et al. 2021). Our results seem to indicate that PBM effects could persist longer than a few minutes. In addition, our comparison between the first and second halves of the session did not reveal any differences between early and late blocks (supplementary material S3). This suggests that the light-induced effects persisted throughout the entire scanning session. A recent small-sample study on the effect of PBM on functional connectivity measured in the prefrontal cortex at rest and during a working memory task, before and 5 days after a single PBM session suggest long-term effects, but the authors measured functional connectivity and not directly haemodynamic responses (O’Connor et al. 2025). Therefore, this question remains to be addressed in further extent.

### PBM induced distinct patterns of effects in young and aged brains

As illustrated on Figure 4, PBM did not appear to elicit widespread changes of BOLD responses across the brain, but rather to have focal effects in several of the task-related regions. In the young group, we found very few differences between PBM and SHAM. In contrast, the aged group showed an increase in BOLD signal from Pre to Post sessions following PBM across several regions. Whole-brain analyses highlighted the involvement of the right ventral premotor region and insula. In addition, further changes were found after restricting the analysis to sensorimotor regions, showing that PBM increased BOLD signal in the left primary motor cortex, right ventral and dorsal premotor cortex, and striatum, to a larger extent when compared to the young group. This indicates that PBM induced focal increases of BOLD responses in some task-related, sensorimotor regions, and this is in line with previous results that have reported increased BOLD signal and functional connectivity after PBM (Dmochowski et al. 2020; Bibb et al. 2025; Yang et al. 2025). In our study, the effect was significant in the aged subjects only, consistent with several preclinical and human studies (Saucedo et al. 2021; Moro et al. 2013; Buendia et al. 2022) suggesting that PBM has a stronger impact on dysfunctional or diseased cells than in healthy ones. For example, Saucedo et al. (2021) used broadband NIRS in aged and young subjects and found that PBM decreased deoxyhemoglobin concentration in both young and aged participants, but increased concentration of cytochrome c oxidase and oxyhemoglobin in only the aged ones. These findings may be related to the fact that ageing and age-related brain diseases are associated with mitochondrial dysfunction (López—Otín et al. 2013) and PBM is thought to improve mitochondrial function and ATP production. Even if this remain to be confirmed, our exploratory analysis suggests that tPBM may affect regions which showed reduced BOLD responses pre-stimulation in aged subjects relatively to the young ones, such as the left primary motor cortex and right striatum. In contrast, no PBM effects was observed in the right primary motor cortex, which showed hyperactivation in aged subjects compared to the young ones. These results suggest that PBM may represent a method for influencing brain metabolism and cerebral haemodynamic.

### PBM did not induce behavioural changes

However, it is worth noting that in our study the increased BOLD response in sensorimotor regions did not result in an improvement of the movement execution rate in the aged participants. In the Young group, the Post-Pre BOLD signal in the left primary motor cortex was positively related to the increase in motor execution rate, regardless of treatment condition. In contrast in the aged group, no such correlation was observed. The reason may be related to the nature of the task which involved many cognitive subprocesses (sequence processing, visuo-motor pairing, movement planification, movement execution). It is possible that different strategies are employed by young and aged subjects, the latter showing less automatised processes and more cognitive ones (Heuninckx et al. 2005). As a result, aged participants would depend more heavily on internally generated motor planning, as reflected by increased baseline activation in supplementary motor and prefrontal regions. In addition, previous studies have also suggested that aged participants rely more on the processing of visual indices and visuo-motor integration to execute this type of tasks (Van Ruitenbeek et al. 2023). This interpretation is consistent with the higher activations observed in occipital and parietal areas in the aged group relatively to the young one (Figure 3).

The fact that the PBM-induced increase in BOLD signal was not reflected in behavioural improvement may also be due to the use of a single-session of PBM that was simply not enough to increase scores at the behavioural level. Investigating the effects of repeated PBM sessions would be important, as positive cumulative effects have been observed with other neurostimulation methods when applied repeatedly. For example, a meta-analysis on the effects of transcranial direct current stimulation over the motor cortex reported significant improvements in motor sequence learning after multiple sessions (3 to 4 days) but not after a single session (Hashemirad et al. 2016). In addition, greater improvements in working memory scores in a N-back task have been showed after seven daily PBM sessions than after a single session (Qu et al. 2022). In light of this, increased BOLD responses in task-related regions could be seen as an early indicator of the changes that accumulate during repeated sessions.

### PBM appeared to act at different levels of the motor control in aged subjects

Our results on laterality of effects are worthy of further discussion. As expected for a right-handed motor task, we obtained activations in both hemispheres with a contralateral dominance in young and aged subjects. However, the PBM-induced effects in the aged subjects were seen mainly on the ipsilateral side, except for the primary motor cortex. Higher ipsilateral activations in motor and premotor areas, as well as increased interhemispheric functional connectivity, have been shown consistently in this type of motor tasks in aged participants compared with younger ones (Turesky et al. 2016; Wang et al. 2019). Such increased activations have been interpreted either by compensation effects, or as the result of a loss of specificity in the neural networks – a dedifferentiation effect.

In young subjects, it has been showed that ipsilateral motor and premotor recruitment increases with task complexity and precision demands (Verstynen et al. 2005). In addition, increased connectivity from right ventral premotor to right primary motor cortex has been proposed to support motor planning and execution when realising complex movements, contributing to the refinement of the motor plans (Bencivenga et al. 2023).

In this context, age-related differences in BOLD responses observed in the present study are consistent with a task-dependent reorganisation of neural activity with aging, as proposed by the HAROLD (Cabeza, 2002; Knights et al. 2021) or CRUNCH hypotheses (Reuter-Lorentz and Cappell 2008). In particular, the CRUNCH model postulates that additional neural recruitment in aging varies as function of task demands and available processing resources. Within this framework, increased baseline BOLD responses may reflect compensatory recruitment of additional neural resources, but may also indicate reduced neural efficiency depending on task demands and available processing capacity. The observed patterns of reduced baseline activation in contralateral motor and striatal regions alongside increased recruitment of supplementary motor, parietal, and occipital areas suggests a redistribution of neural resources toward higher-order visuomotor and control processes in older adults.

In this context, the PBM-related increase of BOLD responses might not be interpreted strictly as beneficial or detrimental, but rather as reflecting a change of neural responsiveness within task-relevant networks. While the complex inter-hemispheric interactions underlying motor control are not yet fully understood, the present findings suggest that PBM may modulate task-related BOLD responses within distributed motor networks in older adults. However, whether such changes reflect improved efficiency, altered resource allocation, or compensatory engagement remains to be determined and likely depends on baseline resources and task demands.

### Limitations

A limitation of this study is that it is a single-blind rather than double-blind design. While double blinding is generally desirable in clinical research studies, its implementation with PBM devices remains challenging due to the practical and technical constraints related with the delivering of active versus sham illumination. Although participants were blinded to the treatment allocation, investigator blinding could not be fully achieved in this study. The primary outcome was nevertheless based on BOLD fMRI signals, that constitute an objective neurophysiological marker that is inherently less susceptible to expectancy and reporting biases than a subjective self-report or behavioural outcome. Consequently, while some observer-related biases cannot be entirely excluded, their potential influence on the main findings should be limited.

## Conclusions

In conclusion, we found that a single session of transcranial PBM with red and near infrared light increased BOLD responses during the execution of finger sequences, in aged subjects, in several regions associated with the performance of the task. Despite the use of a whole-brain device, our findings revealed that tPBM did not elicit global changes across the brain, but rather focal changes restricted to task-related regions over the left primary motor, and higher-order cognitive control areas. Further research should be dedicated to better understand the relative contribution of neural effects and vascular effects. Finally, while these results suggest that tPBM induced neurophysiological changes in the aging brain, additional studies are warranted to clarify the relationship with behavioral outcomes and cognitive functions.

## Data Availability

All data produced in the present study are available upon reasonable request to the authors

## 6. AKNOWLEDGEMENTS

We would like to thank Lilia Langar, Julie Rindone, Marine Mastrosimone for their technical help in the experimentations, Malvina Billères and Marion Coquand-Gandit for their help in the coordination of the project. We also thank Catherine Hamilton and Ron Brown from WellRed for their help in the submission to the ethics. We would like thank the Center for Magnetic Resonance Research from the University of Minnesota for providing the multiband sequence.

This project was funded through a private funding accorded by COVEA foundation to Fonds Clinatec.

