## Supplementary Material S3 for "Transcranial photobiomodulation influences BOLD responses during finger sequence execution: An fMRI Study in young and older adults"

**SUPPLEMENTARY MATERIAL S3. Investigation of treatment effect between early and late blocks.**

To evaluate whether the effects of tPBM found in aged participants were either sustained throughout the entire session or decreased over time, we divided each of the four sessions into two equal parts corresponding to early (first half of the session) and late (second half of the session) blocks. A full-factorial model was then performed, with Timing (Pre, Post), Treatment (PBM, SHAM), and Blocks (Early, Late) as factors.

Supplementary Figure S4A shows whole-brain results for the (Post-Pre)_PBM_ contrast for early (warm colours) and late (green) blocks separately. At an exploratory threshold, the statistical maps suggest that, following tPBM, BOLD signal increased in the right putamen, right dorsal premotor cortex and supplementary area during late blocks while it increased in the right frontal region during early blocks. These differences between early and late blocks did not however, reach statistical significance. Overall, our findings indicated that the effects of tPBM did not differ substantially between the two blocks of the session at the whole brain level.

To investigate further the temporal stability of the effect, we extracted mean contrast estimates from the functional ROIs that showed significant tPBM-induced effects, namely left primary motor, right dorsal and ventral premotor cortex, supplementary motor area and right putamen (Figure S4B-F).

As shown in supplementary Figure S4, no significant Block*Treatment*Timing interaction was detected in any of the regions examined (all p>0.05). In all regions, BOLD signal appeared to increase from Pre to Post PBM condition, irrespective of whether data were extracted from early or late blocks. Taken together, these results indicate that the magnitude of the tPBM-induced effects did not vary across each of the sessions, suggesting that they were maintained throughout the entire session.

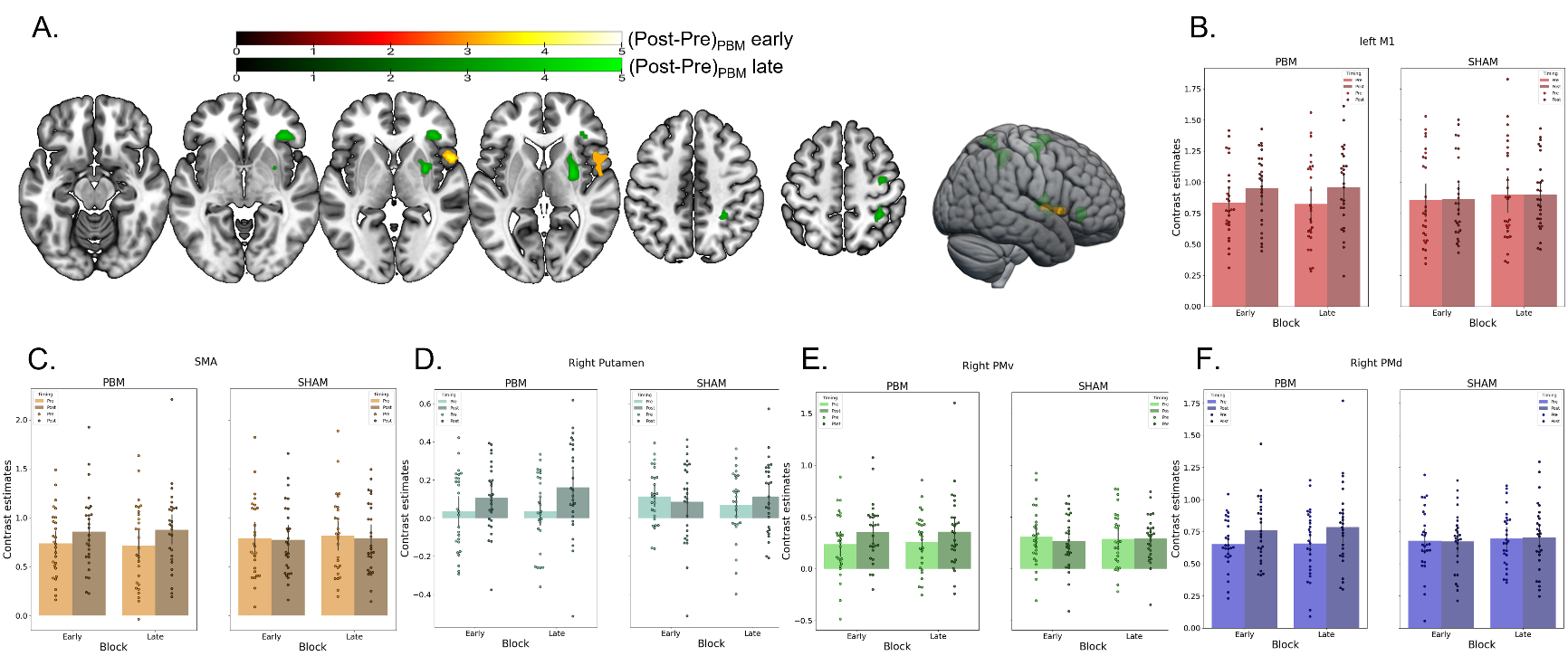

**Figure S3**. **Effect of time on BOLD changes induced by tPBM in the aged group**. **A.** (Post-Pre)_PBM_ contrast displayed on the MNI152 Template, for the early (hot colours) and late (green) blocks. The threshold was set at p<0.001 uncorrected, k≥30 voxels. **B-F.** Mean contrast estimates for the Pre (light colors) and Post (darker colors) sessions, in early and late blocks, and for the PBM (left plots) and SHAM (right plots) treatments. B: left primary motor cortex. C: supplementary motor area. D: right putamen. E: right ventral premotor cortex. F: right dorsal premotor cortex.
