## Supplementary Material S4 for "Transcranial photobiomodulation influences BOLD responses during finger sequence execution: An fMRI Study in young and older adults"

**SUPPLEMENTARY MATERIAL S4. Assessment of motion-related confounds**

To assess potential motion-related confounds, mean framewise displacement (FD) were extracted from the *fmriprep* outputs for each subject and each session. A linear mixed model was then performed to evaluate the effect of Group, Timing and Treatment factors as fixed effects, and Subjects as a random effect.

The results of this analysis are presented in supplementary table S4. A significant main effect of Group was observed, indicating that aged subjects had, in general, greater head motion than young subjects (mean_Young_= 016, S.D.= 0.05; mean_Aged_= 0.26, S.D.= 0.09). In contrast, no main effect of Timing or Treatment were detected, together with no significant interaction observed (all p>0.05).

This suggests that although aged subjects showed greater head movements than young subjects, this did not change across our treatment conditions. Hence, it is most unlikely that differences in head motion contributed to the observed Post-Pre differences between PBM and SHAM treatment.

|  | F-value | p-value |
| --- | --- | --- |
| Group | 26.65 | <0.0001*** |
| Timing | 0.48 | 0.49 |
| Treatment | 0.54 | 0.46 |
| Group*Timing | 0.34 | 0.56 |
| Group*Treatment | 2.12 | 0.15 |
| Timing*Treatment | 0.23 | 0.63 |
| Group*Timing*Treatment | 0.15 | 0.70 |

**Supplementary table S4**. Results of the ANOVA evaluating the effect of Group, Timing, and Treatment on the mean FD.

Further, in order to confirm that the baseline group differences in cerebral activation were not driven by motion differences between the two groups, baseline contrasts young>aged and aged>young were re-evaluated while adding the mean FD as a covariate.

Supplementary Figure S4 presents the results of this analysis. Inclusion of the FD as a covariate yielded findings largely consistent with those obtained in the primary analysis. Specifically, aged participants continued to show reduced activation in left and right primary motor cortex, insula and cerebellum relative to the young ones. In addition, activation was still higher in the paracentral lobule and posterior supplementary motor area, left middle and superior occipital gyrus, and left precuneus.

These results indicate that the baseline difference in brain activation between young and aged participants cannot be readily explained by group differences in head motion.

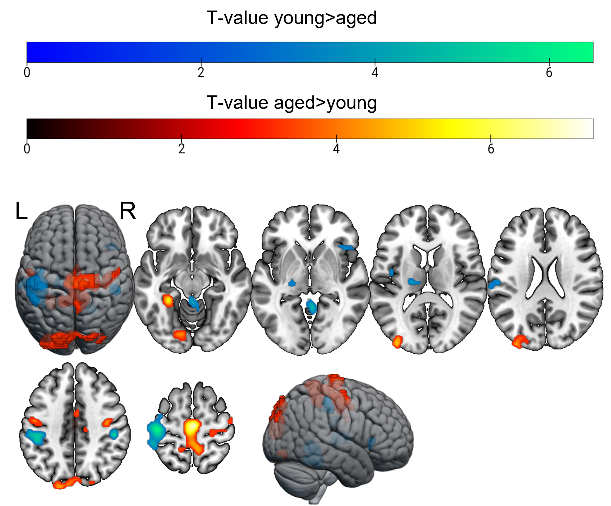

| Contrast | Region Label | MNI coordinates | T-value |
| --- | --- | --- | --- |
| young > aged |  |  |  |
|  | L pre/post central gyr. | [-45 -32 48] | 6.56 |
|  | R postcentral gyr. | [46 -29 44] | 6.03 |
|  | L Thalamus - VPL | [-17 -20 6] | 5.26 |
|  | Cerebellum – vermis 4-5 | [4 -50 -1] | 5.00 |
|  | R cerebellum – 4-5 | [22 -50 -22] | 4.83 |
|  | L postcentral gyr. | [-57 -20 27] | 4.78 |
|  | *L insula *** | *[-42 -8 10]* | *4.14* |
|  | *R inferior frontal gyr. *** | *[43 22 2]* | *3.85* |
| aged > young |  |  |  |
|  | L Paracentral lobule-L/R SMA | [1 -20 55] | 7.34 |
|  | L precuneus | [-14 -41 52] | 5.85 |
|  | L parahippocampal gyrus | [-26 -41 -12] | 5.63 |
|  | L Vis Motor | [-20 -84 52] | 5.58 |
|  | L middle occipital gyrus | [-35 -90 13] | 5.02 |
|  | L superior occipital | [-8 -87 44] | 4.76 |
|  | *L pre/postcentral*** | *[-48 -11 41]* | *4.68* |
|  | *L lingual gyr.-cerebellum *** | *[-14 -81 -12]* | *4.27* |
|  | *R superior occipital gyr.*** | *[31 -84 30]* | *3.85* |

**Figure S4.** **Baseline differences in brain activation between aged and young subjects, with FD added as a covariable in the model**. Left: Brain activation differences displayed on a 3D representation of the MNI152 Template. Statistical map thresholded at p_FWE_<0.05 at the voxel level. Right: Peak localisation and statistics for young>aged and aged>young contrasts, thresholded at p_FWE_<0.05 at the peak level. ***: indicates significant peaks at a lower threshold, p<0.001 uncorrected (peak-level) with an extend threshold k≥30 voxels. L. = left, R. = right. BA: Broadman’s area.
