## Supplementary Material S2 for "Transcranial photobiomodulation influences BOLD responses during finger sequence execution: An fMRI Study in young and older adults"

**Supplementary Material S2. Monte Carlo simulations.**

To evaluate the quantity of energy reaching the cortex, we used Monte Carlo simulation (Prahl et al. 1989). Monte Carlo simulation is a powerful probabilistic solution to simulate light propagation in complex media and has also been used to the brain tissues (Cassano et al. 2019; Yuan et al. 2020; Dole et al. 2024). To take in account the complex geometry of brain tissues, we used meshed-based simulation using the open-source software MMC (Fang and Yan, 2019). Briefly, a T1- and T2-weighted MRI image from a young subject of the CamCan Database (Shafto et al. 2014) was segmented using SimNIBS (Thielscher et al. 2015) in its tissue layers, namely skull, scalp, CSF, grey matter, white matter, air and blood, and then mesh-transformed using Iso2Mesh (Tran et al. 2020). The meshed head contained 237798 nodes and 1 419 136 elements. We used the CamCAN database because of the availability of the T2-weighted image, allowing for a better segmentation of blood vessels. Each of these tissues have their own intrinsic and wavelength-dependent optical properties, which are given by optical coefficients, namely absorption coefficient *µ_a_*, scattering coefficient *µ_s_*_,_ anisotropy factor *g* and refractive index *n*. These optical coefficients were taken from the literature (Bosschaart et al. 2014; Cassano et al. 2019; Dole et al. 2024; Yarolavski et al. 2002) and are described in the following table S1.

|  | 670 nm | | | | 810 nm | | | |
| --- | --- | --- | --- | --- | --- | --- | --- | --- |
|  | µ_a_ | µ_s_ | g | n | µ_a_ | µ_s_ | g | n |
| Air | 0.000001 | 0.10 | 1.00 | 1.00 | 0.000001 | 0.10 | 1.00 | 1.00 |
| White Matter | 0.07 | 40.10 | 0.90 | 1.37 | 0.092 | 38.00 | 0.87 | 1.37 |
| Grey Matter | 0.02 | 8.40 | 0.90 | 1.37 | 0.028 | 7.30 | 0.9 | 1.37 |
| CSF | 0.0004 | 0.30 | 0.90 | 1.37 | 0.0026 | 0.10 | 0.9 | 1.37 |
| Skull / Bone | 0.0208 | 10.81 | 0.89 | 1.37 | 0.011 | 17.45 | 0.89 | 1.37 |
| Soft Tissues | 0.034 | 23.00 | 0.89 | 1.37 | 0.019 | 16.73 | 0.89 | 1.37 |
| Blood | 0.14 | 87.25 | 0.98 | 1.354 | 0.40 | 76.10 | 0.98 | 1.354 |

**Table S1**. Summary of the optical coefficients used for the Monte Carlo simulations. µ_a =_ absorption coefficient; µ_s_ = scattering coefficient; g= anisotropy factor; n = refractive index. Absorption and scattering coefficients are given in /mm^-1^.

A total of 10^7^ photons were launched for each light source, as our previous tests showed that a total of 10^7^ photons ensures good reliability of results using Monte Carlo. At each interaction site, the algorithm calculated the probability of photon being scattered, absorbed, or refracted, as function of the tissue’s optical coefficient; then, the photon was moved and its location updated, and so on, until all photons were absorbed.

In order to register the spatial distribution and orientation of the light source with the MRI image, the 3D locations of each LED were digitised using the Polhemus electromagnetic digitation system (Fastrak, Polhemus Inc., VT, USA). On the helmet, each 670 nm and 810 nm LEDs were positioned side by side, in 40 pairs; for the sake of simplicity, we digitised the mean position between the two LEDs in each pair. The correspondence between digitised LED locations and MRI coordinates were refined after generating a head surface from the T1 MRI image using Brainstorm software (Tadel et al. 2011). As an output, we obtained maps representing the fractional energy deposited per unit of tissue volume (J/mm^3^ per incident J at each source).

Figure S2 shows an example of the maps obtained for each 12-min at 670nm and 12-min at 810nm sessions.

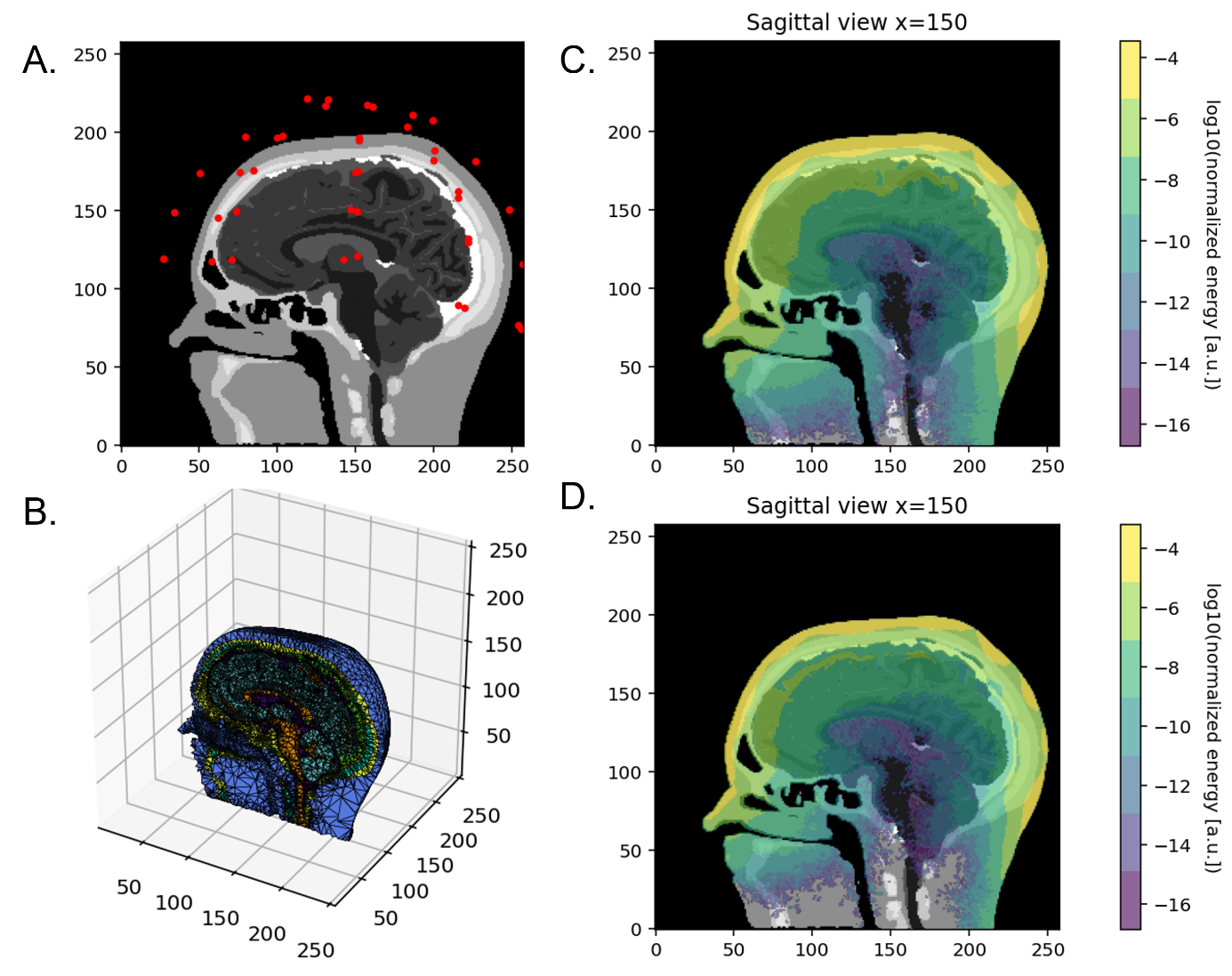

**Figure S2**. A: Localisation of the LED sources superposed on a sagittal view of the brain. B: Illustration of the mesh. C: Normalised energy deposition map (log(J/mm^3^ per joule applied at each source)) for the 810nm wavelength. D: Normalised energy deposition map (log (J/mm^3^) per joule applied at each source) for the 670nm wavelength.

We also extracted and summed the values of normalised energy deposition obtained for each of the nine fMRI ROIs (left/right primary motor cortex, left/right dorsal and ventral premotor areas, left/right putamen and supplementary motor area) for the two wavelengths. By multiplying these normalised values by the output power of each LED and the total session duration, we obtained the total energy deposited in the entire region after the entire 24-min session. The values obtained for each region are presented in Supplementary Table S2.

|  | 670 nm | | | 810 nm | | | Total energy  24-min (J) |
| --- | --- | --- | --- | --- | --- | --- | --- |
| ROI | Sum energy (J) | Mean energy (J/mm^3^) | Max energy  (J/mm^3^) | Sum energy (J) | Mean energy (J/mm^3^) | Max energy  (J/mm^3^) |  |
| Left M1 | 0.014 | 4.88 e-06 | 5.63 e-05 | 0.049 | 1.71 e-05 | 2.05 e-04 | 0.063 |
| Right M1 | 0.014 | 6.73 e-06 | 6.86 e-05 | 0.043 | 2.11 e-05 | 1.99 e-04 | 0.057 |
| Left PMd | 0.016 | 6.19 e-06 | 5.68 e-05 | 0.057 | 2.16 e-05 | 2.14 e-04 | 0.073 |
| Right PMd | 0.046 | 1.91 e-05 | 1.70 e-04 | 0.142 | 5.98 e-05 | 4.63 e-04 | 0.188 |
| Left PMv | 0.029 | 1.38 e-05 | 1.19 e-04 | 0.099 | 4.79 e-05 | 4.05 e-04 | 0.128 |
| Right PMv | 0.023 | 1.68 e-05 | 1.68 e-04 | 0.073 | 5.19 e-05 | 4.70 e-04 | 0.096 |
| SMA | 0.008 | 2.43 e-06 | 1.65 e-05 | 0.025 | 7.72 e-06 | 4.56 e-05 | 0.033 |
| Left Putamen | 1.59 e-05 | 5.32 e-09 | 8.88 e-08 | 3.84 e-05 | 1.29 e-08 | 1.33 e-07 | 5.43 e-05 |
| Right Putamen | 8.12 e-06 | 4.48 e-09 | 1.08 e-07 | 1.56 e-05 | 8.64 e-09 | 1.16 e-07 | 2.37 e-05 |

**Table S2**. Energy deposition values obtained after Monte Carlo simulation, in the nine regions of interests used for fMRI analysis. The sum of energy represents the sum of energy deposited in all the voxels on the region.
